# Parent/caregiver needs during pediatric genome-wide sequencing: a scoping literature review

**DOI:** 10.1101/2025.07.15.25331533

**Authors:** Priyanka Murali, Joon-Ho Yu

## Abstract

**Purpose:** The integration of genome-wide sequencing (GWS), including whole-exome and whole-genome sequencing, has transformed pediatric diagnostics, yet the needs of parents and caregivers during this process remain insufficiently explored. This scoping review aims to synthesize current knowledge on parental and caregiver needs across the GWS process in pediatric settings to inform better clinical practices and support systems.

**Methods:** A scoping review was conducted following PRISMA guidelines. Electronic databases including PubMed, PsycINFO, CINAHL, Embase, and Web of Science were searched, yielding 574 studies, with 47 meeting the inclusion criteria. Data extraction focused on study characteristics, clinical settings, and identified parental needs categorized into pre-test, interim, and post-test periods. Conventional content analysis was used to inductively code and identify themes.

**Results:** Parental needs were categorized into two main themes: (1) Informational needs, encompassing tailored communication, understanding prognosis, logistics, and evolving information, and (2) Emotional support, emphasizing the importance of initial provider interactions and support from healthcare providers and peer groups. Informational and emotional needs were interrelated, impacting parents’ overall experiences. The review highlighted significant gaps during the interim waiting period, with needs largely focused on pre– and post-test periods.

**Conclusion:** Parents navigating the pediatric GWS process require comprehensive informational and emotional support. Effective communication before testing and empathetic follow-up contribute to positive experiences. Addressing gaps at different time throughout the process and fostering continuous provider and peer support can enhance the integration of GWS in pediatric care, improving family-centered outcomes.

## Introduction

Advances in genetic and genomic testing have transformed the landscape of disease diagnosis, treatment, and prevention (Horton & Lucassen, 2019). The development of these technologies has generated new opportunities for improved diagnosis of genetic disorders as well as target treatments in various clinical contexts (Naidoo et al., 2011). Although genetic tests have long been ubiquitous in pediatric settings, these recent advances have changed the landscape of pediatric genetics.

Technologies like whole-exome and whole-genome sequencing (together referred to as genome-wide sequencing or GWS) are being increasingly used for pediatric patients with heterogenous medical presentations, given higher rates of diagnostic yield compared to previously available molecular and cytogenetic testing (Moore & Richer, 2022). However, there are barriers that prevent parents and caregivers of pediatric patients from easily navigating medical and non-medical systems as they go through the process of genetic testing (Dusic et al., 2022). These barriers and gaps are related to a lack of support at different points along the genetic testing process, including differential access, challenges to obtaining informed consent and returning results, as well as gaps in healthcare infrastructure (Boothe et al., 2021; Maiese et al., 2019; Nisselle et al., 2021). As such, the clinical translation of this research requires a deeper understanding of how the implementation of genetic testing technologies is confronted and challenged by systemic problems that challenge adoption of new technologies by patients and their families.

GWS testing is often delineated into pre– and post-test periods which are thought to require different forms of counseling (Elliott, 2020), (McGlynn & Langfelder-Schwind, 2020). The time period preceding testing (pre-test period) begins with an explanation of the method of testing used, the associated risks and benefits, and a discussion about the range of results that can be generated (McGlynn & Langfelder-Schwind, 2020). Then, a sample is collected and there is a period of waiting for results (interim period). In the posttest period, or the time after test results are available, information is shared and the families’ understanding is assessed, expanded, refined, and/or corrected, potentially even creating a medical management and treatment plan (McGlynn & Langfelder-Schwind, 2020). The needs of parents of pediatric patients change as they go through this process, given that both the information and the context in which they receive it typically differ in the pre-test, interim, and post-test periods(Rink & Kuller, 2018).

Studies of parental perspectives and expectations in the context of single gene testing have identified parental needs (Hendriks et al., 2005), (Grosfeld et al., 2000). These studies have identified mitigation of anxiety and a need to better understand current and future impact of genetic testing results among parental needs during the genetic testing process. However, to our knowledge, no review of the current literature has examined the needs of parents and caregivers of pediatric patients as they have navigated the process of GWS testing across different clinical contexts. In addition, parental needs and experiences in different time periods of the GWS process have yet to be explored, including how these needs may be similar or different between time periods. Synthesizing this information may be useful in understanding how to improve parent experiences of GWS and at what time points during the GWS process to target potential interventions. We contend that as GWS becomes increasingly integrated into medical care, failing to understand patient-stakeholder perspectives may hinder effective implementation and exacerbate existing barriers to appropriate genetic and associated follow-up care.

Therefore, we conducted a scoping review of the peer-reviewed academic literature to query the support needs of parents and caregivers of pediatric patients undergoing GWS. The primary aim of this review was to synthesize existing research to better understand these support needs throughout the GWS process. Additionally, we aimed to identify patterns or variations in these needs at different stages of the GWS journey. By doing so, we hope to inform future research and clinical practices that can more effectively address the unique challenges faced by families navigating pediatric GWS.

## Methods

### Scoping review question

The primary research question guiding this scoping review is: “What is the current state of knowledge about the needs of parents and caregivers throughout the process of GWS in pediatric patients?”

To answer this question, we undertook a scoping review of the literature. A scoping review examines the extent, range, and nature, of a particular activity and works to identify gaps in the existing literature (Levac et al., 2010). Scoping reviews are useful for examining emerging evidence when it is still unclear what other, more specific questions can be posed and addressed by a systematic literature review. Given that the state of support needs in this specific context has not fully been examined, a scoping review will be useful to determine the depth and breadth of knowledge regarding this particular topic.

### Search strategies

An initial search of electronic databases and gray literature was conducted, following PRISMA search and screening guidelines for scoping reviews (Tricco et al., 2018). We generated search terms, built search strings, and conducted searches using these strings tailored to specific databases. PubMed, PsycINFO, CINAHL, Embase, and Web of Science databases were searched. Search terms were generated by first identifying concepts that encompass different parts of the question. These concepts included: “pediatrics,” “whole genome sequencing,” “parental or caregiver,” and “attitudes/needs/perspectives.” Relevant MeSH terms were identified based on these concepts of interest. Synonyms to MeSH terms were brainstormed and searched within each database. Some search strings were pre-built for specific databases. Other search strings were built by the first author and research librarian. Starting with MeSH terms, terms for other databases were modified to meet the specifications of the particular database. When terms from one database did not have an equivalent in other databases, those terms were included as free text in the search string. Full search strings for each database are included in the appendix. Titles and abstracts of all articles were searched.

### Inclusion and exclusion criteria

We sought English-language studies focused on understanding the needs, perspectives, and attitudes of parents and caregivers of children who had undergone GWS, defined as either whole exome or whole genome sequencing. To be included, studies had to solicit the perspectives of parents or caregivers of pediatric (aged 0-18 years) patients who had undergone GWS. Studies that included multiple participant groups, such as pediatric patients themselves or clinicians or lay people in addition to parents/caregivers were included if the data for each group could be isolated. Studies must have returned results of GWS to parents/caregivers to be included. Qualitative, quantitative, and mixed methods studies were all included.

### Study selection

Database searches were conducted by the first author on in the following databases: PubMed, PsycINFO, CINAHL, Embase, and Web of Science on May 2^nd^, 2023 and search results imported into Zotero reference management platform. Duplicates were removed prior to uploading the titles and abstracts into Covidence, an online literature review management system. Titles and the abstracts of uploaded articles were screened for inclusion criteria and, if met, a full text review was conducted by the first author.

### Data extraction

The following data were extracted from each included study using an extraction template: (a) bibliographic details (author, year, title, journal, DOI), (b) geographic location (c) study aim, (d) study design, (e) clinical subspecialty in which study was conducted, (f) participant relationship to child, (g) total number of participants in the study, (h) parent/caregiver experiences, expectations, and perceptions, (i) needs preceding, during, and following GWS (if explicitly stated), and (j) stakeholder interactions. The extracted outcomes of this scoping review are parent/caregiver experiences, expectations, and perceptions, and parent/caregiver needs at different points during the process of whole genome sequence.

Data about needs throughout the process of GWS testing was stratified based on whether this was a need preceding sample collection and genome-wide sequencing of the sample, during (in the interim between sample collection and return of results), or following return of GWS results. Extracted data were exported from Covidence into an Excel worksheet.

### Data synthesis and analysis

Each article was reviewed three times to determine if all the data had been extracted and all relevant concepts captured. Excel was then used to calculate frequencies of different categories of extracted data, and descriptive and analytical themes.

### Conventional content analysis

We conducted a conventional content analysis(Hsieh & Shannon, 2005) of identified primary research studies to describe the state of knowledge of parents’ needs throughout the process of pediatric

GWS. As is typical with this approach, codes were developed inductively and sorted in categories based on how they were related and linked.

The extracted data from the needs preceding, during and following GWS categories were imported into Atlas.ti. 23.2.1 for Mac. Initial codes were developed based on a review of the data extracted from the first 10% of articles. The data were then coded line by line and more codes were added as necessary based on the content of the data. The data were then reviewed again to ensure that all relevant concepts had been captured. These codes were then sorted in appropriate descriptive categories and ultimately, broader themes. Specifically, analysis focused on how the codes applied across all need categories were similar or different. These categories and themes were further refined and finalized following discussion between the qualitative research team.

## Results

### Search outcome

The database search returned 574 records. Once duplicates were removed, there were 338 studies remaining for title and abstract screening. Of those, 270 were deemed irrelevant based on the inclusion and exclusion criteria. This left 68 studies to be assessed for full text eligibility. A total of 47 were eligible for inclusion in the final scoping review. **Figure 1** provides a flow diagram following the Preferred Reporting Items for Systematic Reviews and Meta-Analyses (PRISMA) guidelines of the search and screening process.

**Figure 1.**
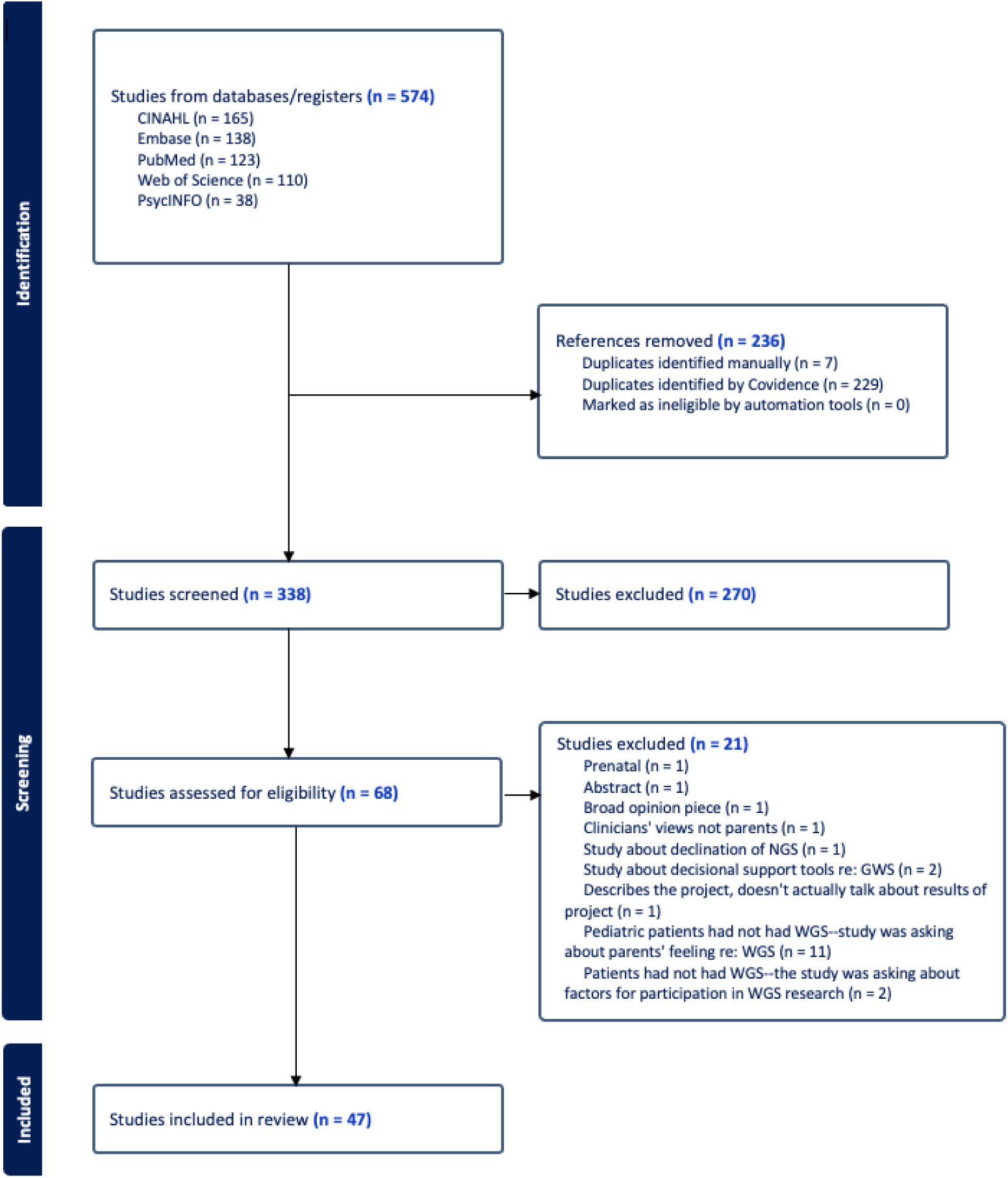
Flow diagram following PRISMA guidelines demonstrating the results of the literature search.

### Study characteristics

The 47 included studies provided data on the needs, experiences, expectations, and perspectives of parents/caregivers of pediatric patients who underwent GWS. Studies were conducted across five countries: the United States, Canada, the United Kingdom, the Netherlands, and Australia. Eight studies focused on GWS in NICU/neonatal settings, seven in oncological settings, six in rare disease settings, four in developmental settings, and four in neurological settings. Four studies involved GWS across multiple clinical settings, and the remaining seven studies addressed various other medical settings, such as cardiac anomalies, congenital malformations, and newborn screening.

Twenty-one studies employed qualitative interviews, 15 utilized surveys, six used mixed methods, and the remainder used other approaches, including but not limited to focus groups. Table 1 provides this information.

**Table 1.** Characteristics of articles.

| Country study was conducted | N (%) |
| --- | --- |
| Australia | 3 (6.38%) |
| Canada | 10 (21.28%) |
| UK | 5 (10.64%) |
| United States | 25 (53.19%) |
| Netherlands | 4 (8.51%) |
| **Total** | **47** |
| Clinical settings of studies | N (%) |
| Developmental | 4 (8.51%) |
| Multiple settings | 4 (8.51%) |
| Neurological | 4 (8.51%) |
| Neonatal/NICU | 8 (17%) |
| Oncological | 7 (14.89%) |
| Other | 14 (29.89%) |
| Rare disease | 6 (12.77%) |
| **Total** | **47** |
| Study design | N (%) |
| Interview – qualitative | 21 (44.68%) |
| Mixed methods | 6 (12.77%) |
| Survey | 15 (31.91%) |
| Other | 5 (10.64%) |
| **Total** | **47** |

### Thematic analysis

Of the 47 articles, 37 mentioned parental needs related to the GWS process. Thematic analysis focused on these 37 articles to identify and classify needs that were preceding, during, and following GWS testing for pediatric patients. We coded more quotations about needs preceding (39%) and following (57%) in contrast to during GWS (3.6%). Identified themes centered informational needs and emotional support, along with related subthemes. **Table 2** provides information on themes and subthemes with supporting quotes.

**Table 2.** Themes, subthemes, and supporting quotes.

| Theme/finding name | Example quote from text |
| --- | --- |
| **Informational Needs** |  |
| Tailoring communication and setting expectations | *Keeping pre-test counseling brief and integrating this with a family systems approach can be very useful in a time-limited situation and to enable decision making for parents who experience decisional conflict or moderate-to-severe anxiety<sup>23</sup>.*<br>*Families being approached for genomic sequencing may benefit from clinicians highlighting the increased probability of a rare or novel gene finding when compared to traditional targeted genetic testing<sup>22</sup>.* |
| Information about logistics, prognosis, and next steps | *Parents across all interviews identified a desire to better understand their child's long-term prognosis as a key motivator to pursue whole-exome sequencing<sup>25</sup>.* |
| Future information changes | *We also found that parents...may want more information on certain topics, such as which genes had been tested and whether reanalysis would be available to them in the future<sup>20</sup>.* 7/15/2025 2:37:00 PM |
| **Emotional impact and support** |  |
| Initial interactions with providers | *...we feel that many of the inequities revealed in our study could be addressed within the current model of genetic counseling, for example with improved 'contracting,' or the dialogue exchanged at the start of a genetic session that establishes the patient-provider relationship<sup>34</sup>.* |
| Support from providers & peer groups | *The importance of parent-to-parent support groups throughout the diagnostic journey was emphasized by parents in both cohorts, suggesting that peer connection may relieve social isolation for parents of children with rare disease<sup>22</sup>* |

#### Informational needs

##### Providers need to tailor communication and set expectations appropriately

An overview of these studies indicated that parents expected clear communication from providers to help them understand the overall GWS process. In the pre-test period, articles frequently emphasized parental reliance on providers to set and shape their expectation (Halley et al., 2022), (Peter et al., 2022), (Lee et al., 2022). This included the need for a clear and easy-to-understand informed consent process to set expectations for testing (Mollison et al., 2020), (Hill et al., 2020), (Hitchcock et al., 2020) and a desire for clarity and acknowledgement that GWS findings might not lead to different treatment or provide different prognostic information (Martinussen et al., 2022). Tailoring pretest counseling conversations was discussed different articles, based both on patient understanding and according to specific clinical scenarios. For example, in NICU settings, parents requested succinct pretest conversations (Wainstein et al., 2022); yet in the post-test period, studies reported overall parental dissatisfaction with the fact that they had received less information than desired (Brett et al., 2019).

##### Parents needs ways to gain information about logistics, prognosis, and next steps

In the pre-test period, some articles noted that a significant motivator for pursuing GWS was the desire to better understand prognosis (Alam et al., 2022). In the post-test period, some articles found that parents also wanted information about logistics. Authors highlighted struggles with accessing medical services, medications, and dealing with insurance reimbursement issues (Halley et al., 2022). Some studies highlighted a need for an intermediary to guide them through the healthcare system (Krabbenborg, Vissers, et al., 2016), especially as the end of a diagnostic odyssey might lead to a new therapeutic journey requiring new care arrangements (Krabbenborg, Schieving, et al., 2016). Beyond clinical care, parents sought therapies and services that often required a genetic diagnosis to gain access (Childerhose et al., 2021). Some articles also reported a need for more information regarding secondary findings (Sanderson et al., 2019) as well as a clear understanding of how results might impact the management of their child’s condition, including planned follow-up, medical and otherwise (Krabbenborg, Schieving, et al., 2016), (Liang et al., 2022), (Stuckey et al., 2015).

##### Guidance about future information changes is needed

Study authors discussed topic exclusively in the post-test period. Study authors discussed parents’ desires to understand how variant interpretations might evolve and how recommendations could change over time, as well as seeking clarity on how the relationship between the clinical genetics team and parents might continue in order to address these desires, (Hill et al., 2020). Some parents wanted explicit information on whom to contact and how to contact them (Krabbenborg, Vissers, et al., 2016), while others preferred periodic outreach from genetics professionals (Krabbenborg, Schieving, et al., 2016).

#### Emotional impact and support

##### Initial interactions with providers are key

In the pre-test period, initial interactions with the clinical team had an impact on parents’ ability to process their feelings about the results in the post-test period (K. C. Li et al., 2016). Furthermore, interactions with providers influenced parents’ perceptions of accessibility and coordination of care; when providers maintained regular contact, parents felt their care was more accessible(Hitchcock et al., 2020). In both the interim period while samples were being analyzed and in the post-test period, parents stated that improved accessibility of the clinic and associated providers improved rapport(X. Li et al., 2019).

##### Support from providers and peer groups is required

Some studies found a need for additional parental support in the pre-test period related to their anxiety about genetic testing (Wainstein et al., 2022) and a desire to alleviate feelings of overwhelm(Luksic et al., 2020). Forms of report were not specified. In the post-text period, articles mentioned a general need to feel emotionally supported by healthcare providers following return of results (Krabbenborg, Schieving, et al., 2016), (Liang et al., 2022). Other articles state that parents preferred that healthcare providers take a more active role in providing psychological support either by directing them towards resources or by making appropriate referrals(Malek et al., 2019), (Werner-Lin et al., 2018), (Wynn et al., 2018). Some articles mentioned the desire for connection with support groups for children with similar diagnoses(Alam et al., 2022), (X. Li et al., 2019). Many studies also indicated that parents wished genetics providers would be the conduit for access to such support groups(Martinussen et al., 2022), (Krabbenborg, Vissers, et al., 2016), (Krabbenborg, Schieving, et al., 2016), (Rosell et al., 2016). Other articles highlighted the importance of acknowledging parental frustration and discussing the possibility of uncertain outcomes in the pretest period(K. C. Li et al., 2016). Discussions regarding return of results often co-occurred with the concept of parental overwhelm and anxiety, typically in post-test periods. Parents stated that receiving a diagnosis did not eliminate negative emotions such as anxiety(Peter et al., 2022).

Notably, very few articles discussed needs in the interim period between sample collection and return of results.

## Discussion

In this scoping review, we examined 47 articles to determine the current state of knowledge about parental and caregiver needs while their children as patients underwent GWS. We found that their needs were interconnected and evolved across different stages of the GWS process. Two overarching themes were identified: 1) Informational needs with the following subthemes: setting expectations and tailoring communication; information about prognosis, logistics, and next steps; and future information changes; 2) Emotional impact and support with subthemes: initial interactions with providers, and support from providers and peers.

This review highlights the fact that needs are varied across different stages but interconnected. In the pre-test setting, parents sought information to understand the types of potential results and the logistical aspects of the GWS process. Prior research suggests that effective communication strategies and realistic pretest counseling are essential for helping families navigate the uncertainty and potential complexity of the GWS process. This emphasizes the importance of communication and suggests a need for more overarching support from healthcare systems themselves as parents navigate the healthcare landscape following genetic testing(Walton et al., n.d.). In the post-test setting, their needs shifted toward understanding the specific results they received, their medical implications, and guidance on how to use or interpret these results in the future. These needs are consistent with those of other studies where parents sought guidance about whether the diagnosis should lead to changes in the daily management of their child and the changing nature of GWS research(Krabbenborg, Schieving, et al., 2016). The review also suggests that the information provided and expectations set during the pre-test period influenced how parents perceived and processed information in the post-test period. This finding underscores the importance of tailoring communication to align with parents’ understanding of genetic information and emotional state in the pre-test setting, as well as setting appropriate expectations, to support better comprehension and decision-making in the post-test period. The extension of benefit from pre-test counseling into the post-test setting, specific to GWS, regardless of diagnostic yield has been seen in previously in the literature(Manickam et al., 2021).

In post-test periods, our findings suggest that parents also need appropriate expectation setting, now focused on both the immediate and long-term future. Parents expressed the need to understand how test results may impact medical care and access to non-medical services. They also sought clarity on how GWS results and associated recommendations may evolve over time with future reanalysis or new data that could impact clinical guidance. Other articles examining post-diagnostic care in rare disease contexts have similar findings. They indicate that genetic conditions can pose a challenge to traditional care management models, given that patients often need follow up care and support from different categories of health professionals, likely from several different medical specialties, as well as by social work providers, all of which requires a level of coordination challenging to organize in most health care systems(Ferrara et al., 2017).

Our findings highlight that initial interactions with healthcare providers were also crucial for establishing rapport and offering emotional support to parents. Parents experienced anxiety and feelings of overwhelm throughout the GWS process. In the pre-test period, this anxiety stemmed from the uncertainty of potential results, while in the post-test period, feelings of overwhelm were linked to the complexity and volume of information provided. This suggests that anxiety persists across both stages of the GWS process and underscores the need for providers to deliver information empathetically to help manage parental anxiety and overwhelm. Furthermore, while parents expressed a desire for information, overburdening parents with information appeared to exacerbate feelings of overwhelm, indicating that a careful balance in communication is essential. Other similar studies report that balancing the amount of information provided does not result in lower parental understanding, and framing information in a way that allows patients to leverage their understanding can allow them to gain agency in their medical care(Joseph et al., 2019).

This review also emphasizes the importance of tailoring communication to match parents and caregivers’ level of understanding and emotional state. This highlights the interconnected nature of informational and emotional needs, as meeting informational needs can directly influence emotional well-being. Parents especially valued and emphasized feeling cared for, having continuous relationships with clinicians, receiving empathetic communication, being kept informed while awaiting genetic test results, being connected with informational and psychosocial resources following results disclosure and follow-up(Crellin et al., 2023). This particular sentiment is echoed in other literature, where framing information in a way that enabled patients to leverage their understanding and gain agency over their medical decision-making also had psychosocial benefit(Joseph et al., 2019). Effectively addressing patients’ informational needs helps them feel supported throughout the process, and establishing good rapport early on further enhances their sense of emotional support.

Parents sought support not only from healthcare providers but also from peer support groups, suggesting that they benefit from diverse perspectives beyond just the medical viewpoint. This points to the fact that parents gained support from a sense of solidarity through interactions with peers who experienced similar medical challenges. This is seen in other clinical genetics literature where parents recruited through peer support groups have emphasized the value of social and informational resources provided by these groups(Riggan et al., 2021), (Nelson Goff et al., 2013). Notably, our review indicates that parents rely on healthcare providers to facilitate connections to resources outside the clinical setting, such as peer support networks. Thus, healthcare providers remain integral, either as direct providers of emotional support or as conduits to other supportive resources for parents.

Additionally, much of the literature appears to focus on needs in the pre-test and post-test periods. There are very limited studies that are examining needs during the interim period between sample collection and return of GWS results, and as such, there is limited data on needs during this period.

Lastly, although several articles noted that parents required support at specific points during the GWS process, the specific nature of these needs was not articulated. This highlights a significant knowledge gap in the literature regarding parental needs in GWS. Addressing these identified gaps presents an opportunity for future research to further explore and define these specific support needs. Future studies may also benefit from directly engaging with parents of children who have undergone GWS, to gain insights into their experiences and assess how their needs align with or diverge from those outlined in this review.

## Limitations

The findings of this scoping review should be considered in light of several limitations. Included studies were conducted primarily in high-income countries which may limit the generalizability of these findings to diverse cultural and socioeconomic contexts. Studies with negative or inconclusive results, or those conducted in lower resourced settings may be underrepresented in the literature due to publication bias. This has the potential to skew the findings toward more well-supported themes, potentially overlooking important challenges faced in less studied or less well-resourced environments. Additionally, the inclusion criteria for this review were limited to peer-reviewed articles, which may miss important insights from gray literature. Despite these limitations, this scoping review highlights what is known in the literature and sheds light on what could be further explored with regard to parental needs in GWS.

## Conclusion

In conclusion, this study identified informational and emotional support needs that spanned across different points of the GWS process in various clinical settings. The examination of these needs through contiguous points in the GWS process shed light on how informational and support needs evolve over the course of the process and how they are interconnected across time and through healthcare systems. By understanding these needs and how they arise, providers who are involved in different parts of the GWS process can better support families through the complex and often overwhelming process of pediatric genome-wide sequencing, ostensibly improving the implementation of GWS into pediatric care.

## Data availability

Data available on request

## Ethics approval statement

This scoping review was in accordance with ethical research principles, utilizing only publicly available published data from peer-reviewed sources. As such, no informed consent or ethical approval was required, and the study did not involve direct interaction with human participants or collection of personal identifiable data.

## Patient consent statement

N/a given this is a scoping review

## Permission to reproduce material from other sources

N/a

## Clinical trial registration

N/a

## Author contributions

Priyanka Murali – Conceptualization; Data curation; Formal analysis; Investigation; Methodology; Project administration; Supervision; Writing – original draft; Writing – review and editing.; Joon-Ho Yu – Conceptualization; Methodology; Resources; Supervision; Writing – review and editing.

## Acknowledgements

This was conducted as a part of a dissertation project.

