## Supplemental Information for "Parent/caregiver needs during pediatric genome-wide sequencing: a scoping literature review"

**Supplemental information - Complete search strings by database**

PubMed

(Infant[Mesh] OR Child[Mesh] OR Adolescent[Mesh] OR Pediatrics[Mesh] OR Infan*[tiab] OR newborn*[tiab] OR "new-born*"[tiab] OR prematur*[tiab] OR preterm*[tiab] OR perinat*[tiab] OR neonat*[tiab] OR baby*[tiab] OR babies[tiab] OR toddler*[tiab] OR minors[tiab] OR minors*[tiab] OR boy[tiab] OR boys[tiab] OR boyhood[tiab] OR girl*[tiab] OR kid[tiab] OR kids[tiab] OR child*[tiab] OR schoolchild*[tiab] OR "school-age*"[tiab] OR adolescen*[tiab] OR juvenil*[tiab] OR youth*[tiab] OR teen*[tiab] OR "under-age*"[tiab] OR pubescen*[tiab] OR pediatric*[tiab] OR paediatric*[tiab] OR peadiatric*[tiab])

**AND**

("Whole Genome Sequencing"[MeSH Terms] OR "Exome sequencing"[MeSH Terms] OR "genome sequenc*"  OR “genomic sequenc*” OR "exome sequenc*" OR “genetic sequenc*" OR “gene panel sequenc*” OR “genetic screening”)

**AND**

("parent attitudes"[tiab:~2] OR "parent's attitudes"[tiab:~2] OR "parents' attitudes"[tiab:~2] OR "parental attitudes" OR

"parent perspective"[tiab:~2] OR "parent's perspective"[tiab:~2] OR "parents' perspective"[tiab:~2] OR "parental perspective"[tiab:~2] OR

"parent perspectives"[tiab:~2] OR "parent's perspectives"[tiab:~2] OR "parents' perspectives"[tiab:~2] OR "parental perspectives"[tiab:~2] OR

"parent needs"[tiab:~2] OR "parent's needs"[tiab:~2] OR "parents' needs"[tiab:~2] OR "parental needs"[tiab:~2] OR

"parent knowledge"[tiab:~2] OR "parent's knowledge"[tiab:~2] OR "parents' knowledge"[tiab:~2] OR "parental knowledge"[tiab:~2] OR

"parent experiences"[tiab:~2] OR "parent's experiences"[tiab:~2] OR "parents' experiences"[tiab:~2] OR "parental experiences"[tiab:~2] OR "parent choice"[tiab:~2] OR "parent's choice"[tiab:~2] OR "parents' choice"[tiab:~2] OR "parental choice"[tiab:~2] OR "parent choices"[tiab:~2] OR "parent's choices"[tiab:~2] OR "parents' choices"[tiab:~2] OR "parental choices"[tiab:~2] OR "parent opinions "[tiab:~2] OR "parent's opinions "[tiab:~2] OR "parents' opinions "[tiab:~2] OR "parental opinions "[tiab:~2] OR "parent preference"[tiab:~2] OR "parent's preference"[tiab:~2] OR "parents' preference"[tiab:~2] OR "parental preference"[tiab:~2] OR "parent preferences"[tiab:~2] OR "parent's preferences"[tiab:~2] OR "parents' preferences"[tiab:~2] OR "parental preferences"[tiab:~2] OR "parent concerns "[tiab:~2] OR "parent's concerns"[tiab:~2] OR "parents' concern "[tiab:~2] OR "parental concerns"[tiab:~2] OR

“parent belief"[tiab:~2] OR "parent's belief"[tiab:~2] OR "parents' belief"[tiab:~2] OR "parental belief"[tiab:~2] OR

“parent beliefs"[tiab:~2] OR "parent's beliefs"[tiab:~2] OR "parents' beliefs"[tiab:~2] OR "parental beliefs"[tiab:~2] OR

"parent desire"[tiab:~2] OR "parent's desire"[tiab:~2] OR "parents' desire"[tiab:~2] OR "parental desire"[tiab:~2] OR

"parent desires"[tiab:~2] OR "parent's desires"[tiab:~2] OR "parents' desires"[tiab:~2] OR "parental desires"[tiab:~2] OR

"parent perception"[tiab:~2] OR "parent's perception"[tiab:~2] OR "parents' perception"[tiab:~2] OR "parental perception"[tiab:~2] OR

"parent perceptions"[tiab:~2] OR "parent's perceptions"[tiab:~2] OR "parents' perceptions"[tiab:~2] OR "parental perceptions"[tiab:~2] OR

(Parents[Mesh] AND (“Health knowledge, attitudes, practice”[Mesh] OR Attitude[Mesh])))

Web of Science

(Infan* OR newborn* OR "new-born*" OR prematur* OR preterm* OR perinat* OR neonat* OR baby* OR babies OR toddler* OR minors OR minors* OR boy OR boys OR boyhood OR girl* OR kid OR kids OR child* OR schoolchild* OR "school-age*" OR adolescen* OR juvenil* OR youth* OR teen* OR "under-age*" OR pubescen* OR pediatric* OR paediatric* OR peadiatric*)

**AND**

(“genome sequenc*” OR “genomic sequenc*” OR "exome sequenc*" OR “genetic sequenc*" OR “gene panel sequenc*” OR “genetic screening”)

**AND**

(parent* NEAR/1 (attitudes OR perspective* OR needs OR knowledge OR experiences OR choice* OR opinions OR preference* OR belief* OR desire* OR perception*))

CINAHL (EBSCO)

**(**(MH "Child+") OR (MH "Infant+") OR (MH "Adolescence+") OR (MH "Pediatrics+") OR

TI(Infan* OR newborn* OR "new-born*" OR prematur* OR preterm* OR perinat* OR neonat* OR baby* OR babies OR toddler* OR minors OR minors* OR boy OR boys OR boyhood OR girl* OR kid OR kids OR child* OR schoolchild* OR "school-age*" OR adolescen* OR juvenil* OR youth* OR teen* OR "under-age*" OR pubescen* OR pediatric* OR paediatric* OR peadiatric*) OR

AB(Infan* OR newborn* OR "new-born*" OR prematur* OR preterm* OR perinat* OR neonat* OR baby* OR babies OR toddler* OR minors OR minors* OR boy OR boys OR boyhood OR girl* OR kid OR kids OR child* OR schoolchild* OR "school-age*" OR adolescen* OR juvenil* OR youth* OR teen* OR "under-age*" OR pubescen* OR pediatric* OR paediatric* OR peadiatric*)**)**

 AND

**(**(MH "Genetic Screening+") OR

TI(“genome sequenc*” OR “genomic sequenc*” OR "exome sequenc*" OR “genetic sequenc*" OR “gene panel sequenc*” OR “genetic screening”)

OR

AB(“genome sequenc*” OR “genomic sequenc*” OR "exome sequenc*" OR “genetic sequenc*" OR “gene panel sequenc*” OR “genetic screening”)**)**

AND

**(**

(((MH "Attitude+") OR (MH "Health Knowledge")) AND (MH "Parents+")) OR

TI(parent* N1 (attitudes OR perspective* OR needs OR knowledge OR experiences OR choice* OR opinions OR preference* OR belief* OR desire* OR perception*))

**OR**

AB(parent* N1 (attitudes OR perspective* OR needs OR knowledge OR experiences OR choice* OR opinions OR preference* OR belief* OR desire* OR perception*))

**)**

Psycinfo (EBSCO)

**(**

AG(Childhood OR Adolescence)

OR

MA(“Infant” OR “Child” OR “Adolescent” OR “Pediatrics”)

 OR

KW(Infan* OR newborn* OR "new-born*" OR prematur* OR preterm* OR perinat* OR neonat* OR baby* OR babies OR toddler* OR minors OR minors* OR boy OR boys OR boyhood OR girl* OR kid OR kids OR child* OR schoolchild* OR "school-age*" OR adolescen* OR juvenil* OR youth* OR teen* OR "under-age*" OR pubescen* OR pediatric* OR paediatric* OR peadiatric*)

**OR**

TI(Infan* OR newborn* OR "new-born*" OR prematur* OR preterm* OR perinat* OR neonat* OR baby* OR babies OR toddler* OR minors OR minors* OR boy OR boys OR boyhood OR girl* OR kid OR kids OR child* OR schoolchild* OR "school-age*" OR adolescen* OR juvenil* OR youth* OR teen* OR "under-age*" OR pubescen* OR pediatric* OR paediatric* OR peadiatric*)

**OR**

AB(Infan* OR newborn* OR "new-born*" OR prematur* OR preterm* OR perinat* OR neonat* OR baby* OR babies OR toddler* OR minors OR minors* OR boy OR boys OR boyhood OR girl* OR kid OR kids OR child* OR schoolchild* OR "school-age*" OR adolescen* OR juvenil* OR youth* OR teen* OR "under-age*" OR pubescen* OR pediatric* OR paediatric* OR peadiatric*)

**)**

**AND**

**(**

DE ("Genomic Sequencing")

OR

MA ("Whole Genome Sequencing" OR "Exome sequencing")

OR

KW(“genome sequenc*” OR “genomic sequenc*” OR "exome sequenc*" OR “genetic sequenc*" OR “gene panel sequenc*” OR “genetic screening”)

OR

TI(“genome sequenc*” OR “genomic sequenc*” OR "exome sequenc*" OR “genetic sequenc*" OR “gene panel sequenc*” OR “genetic screening”)

**OR**

AB(“genome sequenc*” OR “genomic sequenc*” OR "exome sequenc*" OR “genetic sequenc*" OR “gene panel sequenc*” OR “genetic screening”)**)**

**AND**

**(**

((DE "Parents" OR DE "Adoptive Parents" OR DE "Expectant Parents" OR DE "Fathers" OR DE "Foster Parents" OR DE "Homosexual Parents" OR DE "Mothers" OR DE "Parental Characteristics" OR DE "Single Parents" OR DE "Stepparents" OR DE "Surrogate Parents (Humans)") AND (DE "Parental Attitudes" OR DE "Parental Expectations" OR DE "Health Attitudes" OR DE "Attitudes" OR DE "Health Behavior" OR DE "Health Knowledge"))

OR

MA ((Parents AND (“Health knowledge, attitudes, practice” OR Attitude))

OR

KW(parent* N1 (attitudes OR perspective* OR needs OR knowledge OR experiences OR choice* OR opinions OR preference* OR belief* OR desire* OR perception*))

**OR**

TI(parent* N1 (attitudes OR perspective* OR needs OR knowledge OR experiences OR choice* OR opinions OR preference* OR belief* OR desire* OR perception*))

**OR**

AB(parent* N1 (attitudes OR perspective* OR needs OR knowledge OR experiences OR choice* OR opinions OR preference* OR belief* OR desire* OR perception*))

**)**

**Embase (Elsevier)**

**(**'adolescent'/exp OR 'child'/exp OR 'infant'/exp OR 'pediatrics'/exp

OR

(Infan* OR newborn* OR "new-born*" OR prematur* OR preterm* OR perinat* OR neonat* OR baby* OR babies OR toddler* OR minors OR minors* OR boy OR boys OR boyhood OR girl* OR kid OR kids OR child* OR schoolchild* OR "school-age*" OR adolescen* OR juvenil* OR youth* OR teen* OR "under-age*" OR pubescen* OR pediatric* OR paediatric* OR peadiatric*):TI,AB,KW**)**

**AND**

('whole exome sequencing'/exp OR 'whole genome sequencing'/de

OR

(“genome sequenc*” OR “genomic sequenc*” OR "exome sequenc*" OR “genetic sequenc*" OR “gene panel sequenc*” OR “genetic screening”):TI,AB,KW**)**

**AND**

**((**'parent'/exp AND ('attitude to health'/exp OR 'attitude'/exp))

OR
(parent* NEAR/2 (attitudes OR perspective* OR needs OR knowledge OR experiences OR choice* OR opinions OR preference* OR belief* OR desire* OR perception*)):TI,AB,KW**)**
